# ASSOCIATIONS OF AGE, BODY MASS INDEX, AND BIOCHEMICAL PARAMETERS WITH BRAIN MORPHOLOGY IN PATIENTS WITH ANOREXIA NERVOSA

**DOI:** 10.1101/2020.06.30.20143362

**Authors:** Lasse Bang, Christian K. Tamnes, Linn B. Norbom, Rut A. Thomassen, Jill S. Holm, Laila H. Skotte, Petur B. Juliusson, Magnus Mejlænder-Evjensvold, Øyvind Rø

**Affiliations:** Regional Department for Eating Disorders, Division of Mental Health and Addiction, Oslo University Hospital, Oslo, Norway; PROMENTA Research Center, Department of Psychology, University of Oslo, Oslo, Norway; NORMENT, Institute of Clinical Medicine, University of Oslo, Oslo, Norway; Department of Psychiatric Research, Diakonhjemmet Hospital, Oslo, Norway; Department of Paediatric Medicine, Division of Paediatric and Adolescent Medicine, Oslo University Hospital, Oslo, Norway; Department of Clinical Neurosciences for Children, Division of Paediatric and Adolescent Medicine, Oslo University Hospital, Oslo, Norway; Department of Clinical Science, University of Bergen, Bergen, Norway; Department of Health Registries, Norwegian Institute of Public Health, Bergen, Norway; Department of Paediatrics, Haukeland University Hospital, Bergen, Norway; Division of Radiology and Nuclear Medicine, Department of Radiology, Oslo University Hospital, Oslo, Norway; Division of Mental Health and Addiction, Institute of Clinical Medicine, University of Oslo, Oslo, Norway

**Keywords:** Aging, Anorexia nervosa, Biochemistry, Body mass index, Brain, Cerebral cortex, Growth and development, Feeding and eating disorders, Gray matter, Magnetic resonance imaging

## Abstract

**INTRODUCTION:** Accumulating evidence shows that patients with anorexia nervosa (AN) have globally reduced brain mass, including lower cortical volume and thickness, which largely normalizes following weight restoration. The dynamic underlying mechanisms for these processes are unknown, and how age and severity of emaciation are associated with brain morphology in AN is poorly understood. We investigated associations of age, body mass index (BMI) and biochemical parameters with brain morphology in a large sample of patients in treatment. METHODS: We included 85 patients (94% female) aged 12-48 (mean = 23) years with clinical and quality controlled magnetic resonance imaging (MRI) data. T1-weighted MRI images, clinical characteristics, and biochemical parameters were retrospectively collected from hospital records. Brain morphology was measured using FreeSurfer, and associations were investigated using regression models and correlations. RESULTS: Controlling for BMI, age showed significant associations with brain morphology generally concordant with known typical brain developmental patterns. Controlling for age, BMI showed significant positive associations with cortical volume and thickness. There were no significant interaction effects between age and BMI. None of the biochemical parameters correlated significantly with brain morphology.

**DISCUSSION:** Our findings suggest the presence of normal neurodevelopmental patterns in AN and highlight the value of considering age-related effects on brain morphology. Importantly, we showed that severity of emaciation is related to brain morphology reductions, underscoring the importance of weight restoration. More studies are needed to shed light on potential biochemical mechanisms associated with brain alterations in AN.

## 1. INTRODUCTION

Accumulating evidence shows that patients with anorexia nervosa (AN) have globally reduced gray matter (GM) and white matter (WM) brain volumes compared to healthy individuals (Seitz, Herpertz-Dahlmann, & Konrad, 2016; Titova, Hjorth, Schioth, & Brooks, 2013; Van den Eynde et al., 2012). The GM reductions comprise both cortical and subcortical tissues, and specifically thinning of the cerebral cortex (Bär, de la Cruz, Berger, Schultz, & Wagner, 2015; King et al., 2015; Miles, Voineskos, French, & Kaplan, 2018; Nickel et al., 2018). Some studies have also found greater cortical volume and thickness in local regions (Boghi et al., 2011; Brooks et al., 2011; Frank, Shott, Hagman, & Mittal, 2013; Lavagnino et al., 2018).

Evidence from longitudinal studies indicates that AN-related brain alterations largely normalize during weight restoration treatment (Bernardoni et al., 2016; Bomba et al., 2015; Kaufmann et al., 2020; Mainz, Schulte-Rüther, Fink, Herpertz-Dahlmann, & Konrad, 2012). Accordingly, recovered patients with normal body weights typically show no significant differences in gross brain morphology compared to healthy individuals (Bang, Rø, & Endestad, 2016; Bernardoni et al., 2016; Lázaro et al., 2013; Wagner et al., 2006), although some studies report persistent small local alterations (Castro-Fornieles et al., 2019; Frank et al., 2013; Lavagnino et al., 2018). Collectively, these findings suggest the alterations in brain morphology are secondary to emaciation, and largely reversible with weight restoration.

The underlying neurobiological mechanisms of the dynamic brain morphology alterations in AN remain unknown. Several hypotheses have been proposed, including dehydration, loss of neuronal or glial cells, and hormonal changes (King, Frank, Thompson, & Ehrlich, 2018). Few studies have investigated associations between biochemical/hormonal parameters and brain morphology, which could shed light on mechanistic relations. Preliminary evidence supports a relationship between brain morphology and hormonal levels, including cortisol (Castro-Fornieles et al., 2009; Mainz et al., 2012) and follicle-stimulating hormone (Mainz et al., 2012). In contrast, two previous studies considered serum albumin and hydration indices as measures of dehydration and fluid shifts, but found that these were within normal range and thus unlikely to contribute to brain alterations (Bernardoni et al., 2016; King et al., 2015).

The extent to which severity of emaciation and age are related to brain morphology alterations in AN is poorly understood. While some studies report positive associations between body mass index (BMI, kg/m^2^) and cortical volume and thickness (Lavagnino et al., 2016; Nickel et al., 2018), others have not found similar relations (Curzio et al., 2020; Fuglset et al., 2016; King et al., 2015). It is therefore unclear whether increased AN severity is related to greater brain morphology alterations. Moreover, age and neurodevelopment constitutes an additional factor that may influence the extent of brain morphology alterations among patients, but few studies have investigated this. It is well-documented that normal neurodevelopment from childhood to adulthood involves GM volume decreases, cortical volume and thickness decreases, and WM volume increases with increasing age (Lebel & Beaulieu, 2011; Mills et al., 2016; Tamnes et al., 2017). A previous study (King et al., 2015) reported an absence of such expected age-related changes in cortical thickness among patients with AN, indicating disrupted neurodevelopment. Similarly, one review concluded that the reductions in brain mass appear to be larger among adolescent as opposed to adult patients with AN (Seitz et al., 2016), raising concerns about the influence of emaciation on neurodevelopment. It was also recently reported that restoration of brain morphology during treatment is age dependent; with younger patients recuperating faster than older patients (Kaufmann et al., 2020). Thus, exactly how age influences brain morphology in AN and whether neurodevelopment is disrupted is unclear. One possibility is that age moderates the relationship between BMI and brain morphology, such that low body weight is associated with greater brain mass reductions among younger as compared to older patients. Studies attempting to untangle the complex relationships between age, BMI, and brain morphology are thus critically needed.

The current study investigated the relationship between brain morphology and clinical characteristics in a large sample of patients with AN receiving weight restoration treatment. Our primary aim was to estimate the associations of age and BMI with brain morphology. We hypothesized that age would exhibit negative associations with cortical volumes and thickness and positive associations with WM volumes, in line with known typical neurodevelopmental patterns. We hypothesized that BMI would exhibit positive associations with brain morphology, in line with prior studies showing normalization of brain tissues following weight restoration. Additionally, we hypothesized an interaction effect between age and BMI, which would indicate the presence of a moderating effect of age on the association between BMI and brain morphology. A secondary aim was to explore potential biochemical correlates of brain morphology in order to elucidate mechanisms underlying brain alterations. As these analyses were considered exploratory, we had no formal hypotheses.

## 2. METHODS

### 2.1 Participants

The final sample consisted of 85 patients (out of 212 eligible), for whom both clinical information and quality controlled neuroimaging data were available. Patients had a primary AN diagnosis and were admitted to one of two specialized treatment units at Oslo University Hospital (Norway), between 2007 and 2018. Both units provide in- and out-patient treatment to patients with eating disorders, with weight-rehabilitation as a primary goal of treatment. One unit admits patients of all ages, and the other admits patients under the age of 18. Patients with AN who are admitted to these units are routinely referred to a clinical magnetic resonance imaging (MRI) examination of their brain, to rule out organic causes of their disordered eating. The timing of the MRI examination varies, but typically occurs about one month after admission (due to medically unstable patients or wait-list for the MRI scanner). Patients diagnosed with AN who had completed an MRI examination were identified retrospectively using hospital records, resulting in 212 eligible patients. The study was approved by the Regional Committee for Medical and Health Research Ethics (#2017/716), and given an exempt from the normal requirement of informed consent.

### 2.2 Clinical information

Clinical information was acquired from patient records, and included details about the patients and their treatment. Information regarding BMI was extracted and converted to standard deviation scores (SDS, adjusted for age and gender) using national Norwegian references (Júlíusson et al., 2013). Patients older than 19 years were assigned an age of 19 years for calculating BMI SDS.

Information on biochemical parameters measured by clinical lab-analyses of venous blood samples was collected from patient records. Specific biochemical parameters were missing for many patients (i.e. they were not collected at the time of the MRI examination), so we only included parameters that were available for 25 patients or more. These included levels of electrolytes (calcium, sodium, potassium, chloride, phosphate, bicarbonate), albumin, base-excess, partial pressure of carbon dioxide (pCO_2_), C-reactive protein (CRP), erythrocyte volume fraction (EVF), lactate dehydrogenase (LD), pH, and hemoglobin. Analyses of blood samples were performed as part of routine clinical evaluation at Oslo University Hospital.

As height, weight, biochemical parameters, and MRI data were originally collected for clinical purposes, they were not necessarily collected on the same day. We only included height/weight measurements that were performed within 31 days of the MRI examination, and biochemical parameters that were performed within 7 days. Patients were weighed on average four days before the MRI examination, and biochemical parameters were collected on average 0.11 days before the MRI examination (see Table 1).

**Table 1.** Clinical characteristics.

|  | *Mean ± SD* | *Median (range)* | *n* |
| --- | --- | --- | --- |
| **Age** | 23.43 ± 8.04 | 21.42 (12.92; 48.17) | 85 |
| **Admission BMI** | 15.04 ± 2.03 | 14.75 (10.16; 19.93) | 84 |
| **Admission BMI SDS** | -3.83 ± 1.79 | -3.58 (-9.70; -0.72) | 83 |
| **BMI at MRI** | 15.93 ± 2.08 | 15.64 (9.71; 20.73) | 80 |
| **BMI SDS at MRI** | -3.16 ± 1.69 | -3.06 (-10.61; -0.38) | 80 |
| **Days from admission to MRI** | 44.73 ± 49.19 | 31.00 (1; 308) | 85 |
| **Days between weighing and MRI** | 4.17 ± 8.36 | 1.00 (-14; 31)<sup>†</sup> | 80 |
| **Days between blood sample and MRI** | 0.11 ± 1.67 | 0.00 (-7; 3)<sup>†</sup> | 76 |
|  | *Count (%)* |  | *n* |
| **Gender** | 80 (94%) Female |  | 85 |
| **Inpatient or outpatient admission?** | 77 (91%) Inpatient |  | 85 |
| **Patient committed involuntarily?** | 18 (21%) Yes |  | 85 |
| **On psychoactive medication?** | 22 (26%) Yes |  | 85 |

Note: <sup>†</sup> Negative values indicate weight/blood sample was collected after the MRI examination.
Abbreviations: BMI: Body mass index (kg/m<sup>2</sup>); BMI SDS: Body mass index standard deviation score; MRI: Magnetic resonance imaging; SD: Standard deviation.

### 2.3 MRI acquisition

Neuroimaging of the final sample was attained on eight separate whole body scanners from GE medical systems (one 3T scanner), Siemens (four 1.5T scanners), and Philips Medical Systems (two 1.5T and one 3T scanners). The specific sequences within each scanner varied, as they were based on clinical protocols. See Supplemental Table S1 and S2 for an overview of the various MRI scanners and sequences and brain morphology estimates for each scanner, respectively. Supplemental Figure S1-3 presents visual depictions of age distributions, BMI, and brain morphology estimates across scanners.

### 2.4 MRI analysis and quality control

T1-weighted datasets were processed using the automatic software suite FreeSurfer 6.0, documented and freely available online (http://surfer.nmr.mgh.harvard.edu/). The technical details of the procedures have previously been described in detail (Dale, Fischl, & Sereno, 1999; Fischl, 2012; Fischl et al., 2002; Fischl, Sereno, & Dale, 1999). In short, FreeSurfer performs volumetric segmentations and cortical surface reconstructions, including the “pial” (gray/cerebrospinal fluid boundary) and “white” (gray/white matter boundary) surface. Cortical thickness was computed as the shortest vertex-wise distance between the white and pial surface, while cortical surface area, based on the white surface, was computed by the amount of vertex-wise contraction and expansion required to fit a common template (fsaverage). Acknowledging the limitations in pooling clinical neuroimaging data from several scanners, we focused on global brain morphology measures; including cortical volume, cortical thickness, cortical surface area, total subcortical volumes, cerebral WM volume, cerebellar GM volume, cerebellar WM volume, and ventricular volumes (the combined volume of the lateral ventricles, the inferior lateral ventricles and the 3^rd^ ventricle). All MRI variables were summed (or averaged for cortical thickness) across hemispheres.

As the MRI data were collected in a clinical context, the data quality varied extensively. FreeSurfer was only able to successfully process 147 of the total 212 eligible MR-images. The resulting 147 FreeSurfer processed scans were subjected to both manual and automatic quality control. First, all images and segmentations were visually inspected post-processing by a trained operator (CKT) for accuracy, and rated as acceptable or not acceptable. Second, Qoala-T 1.2 (Klapwijk, van de Kamp, van der Meulen, Peters, & Wierenga, 2019) was used to automatically assess the quality of segmented data (see Supplemental Figure S4 for results from Qoala-T quality control), yielding recommendations for inclusion or exclusion. The overlap between the two methods was very high, with the same recommendation for 142 of the 147 scans. In cases of disagreement between the manual and the automatic quality control, a second visual inspection was performed to decide on whether to include or exclude the scan. A total of 56 scans were excluded. Six additional patients were excluded as it was discovered that necessary clinical information was unavailable, yielding a total of 85 patients with quality controlled MRI data for the final sample.

### 2.5 Statistical analyses

Pearson *r* correlations were used to investigate associations between age, BMI, treatment duration and estimated intracranial volume (ICV); and Pearson partial correlations (with ICV as covariate) used to investigate associations of days between weighing and MRI examination with brain morphology measures. T-tests were performed to investigate differences in brain morphology, age, and BMI between patients on psychoactive medications to patients who were not. Alpha-levels for these tests were not adjusted for multiple comparisons, as they were not related to our primary aims.

To investigate associations of age and BMI with brain morphology, linear multiple regression analyses were performed. Age and BMI SDS were entered as independent variables (both mean-centered), and an interaction term between age and BMI SDS was included to explore presence of moderating effects. The model is summarized in the following equation:

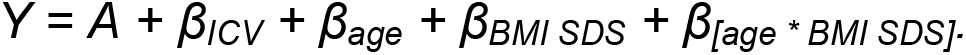

Y represents the measures of brain morphology and A the intercept representing Y at mean age, BMI SDS and ICV. Models were run both with and without ICV as a nuisance variable in order to investigate both relative and absolute individual differences in brain morphology, as recommended in Mills and Tamnes (2014). Regression models of relative brain morphology (models including ICV) are the ones of primary interest in our study. Alpha-levels were corrected for multiple comparisons according to a Bonferroni-Holm adjustment corresponding to number of independent variables of interest (*n* = 6) for each measure of brain morphology. To ease interpretation, we report adjusted *p*-values, with *p* < .05 considered statistically significant.

To investigate associations between biochemical parameters and brain morphology, partial Pearson *r* correlations (controlling for ICV) were performed. Alpha-levels were corrected for multiple comparisons according to a Bonferroni-Holm procedure, corresponding to the number of biochemical parameters of interest (*n* = 15). We report adjusted *p*-values, with *p* < .05 considered statistically significant. All analyses were performed in R version 3.6.1 (R Core Team, 2019).

## 3. RESULTS

### 3.1 Clinical characteristics

Table 1 presents clinical characteristics of the 85 patients included in the analyses. Twenty-two patients (26%) were < 18 years of age, and five were male. Patients had been admitted for an average of 45 days prior to the MRI examination. Between admission and the MRI examination there was a mean weight gain of 2.66 kilograms, corresponding to 0.89 BMI and 0.67 BMI SDS units. There was a small but significant positive correlation between treatment duration and BMI SDS at the time of the MRI (r = .27, p = .02), illustrating that patients gained weight during treatment. Eighty-two percent of patients had a BMI < 18.5 and 76% a BMI SDS corresponding to underweight at the MRI examination. No significant correlation existed between age and BMI SDS (r = −0.17, *p* = .14). Approximately 26% of patients were on psychoactive medications. T-tests showed no significant differences in age, BMI SDS, or any of the brain morphology measures between patients on medication compared to patients who were not (all *p*’s > .18).

Number of days between weighing and the MRI examination did not correlate significantly with the brain morphology variables (*r*’s −.06 to .15, all *p*’s > .19), suggesting the time-lag between weighing and the MRI examination had limited effect on estimates of brain morphology. There was no significant correlation between ICV and BMI SDS (*r* = .15, *p* = .18) or age (*r* = −.03, p = .79).

### 3.2 Associations of age and BMI with brain morphology

#### Associations between age and brain morphology

In the primary regression models adjusted for ICV, age showed significant negative associations with cortical volume, cortical surface area, subcortical volumes, and a significant positive association with cerebral WM volume (Table 2). No other significant associations were evident, although we note that the negative association between age and cortical thickness was nearly significant (*p* = 0.05) prior to alpha-level correction for multiple comparisons. These results imply that increasing age (while controlling for BMI) is associated with tissue specific changes in relative brain morphology in patients with AN, generally in accordance with typical patterns of structural brain development (Mills et al., 2016; Tamnes et al., 2017). In regression models unadjusted for ICV, age showed a significant negative association with cortical volume only (but cortical thickness was significant prior to alpha-level correction).

**Table 2.** Results from regression models.

|  | BMI SDS |  |  | Age |  |  | Interaction BMI SDS * Age |  |  |
| --- | --- | --- | --- | --- | --- | --- | --- | --- | --- |
| Adjusted for ICV (df = 75) | $\beta$ (95% CI) | SE | P | $\beta$ (95% CI) | SE | P | $\beta$ (95% CI) | SE | P |
| Cortical volume | 9491.40 (5581.64; 13645.28) | 2052.50 | < .001* | -2037.94 (-2895.67; -1180.22) | 430.56 | < .001* | -499.70 (-1160.61; 161.22) | 331.77 | .27 |
| Cortical thickness | 0.031 (0.017; 0.044) | 0.007 | < .001* | -0.003 (-0.006; 0.000) | 0.001 | .19 | -0.001 (-0.004; 0.001) | 0.001 | .41 |
| Cortical surface area | 799.49 (-471.22; 2070.20) | 637.88 | .78 | -367.23 (-633.79; -100.67) | 133.81 | .05*<sup>†</sup> | -79.98 (-285.38; 125.42) | 103.11 | .78 |
| Subcortical volumes | -241.77 (-650.23; 166.69) | 205.04 | .83 | -131.05 (-216.74; -45.37) | 43.01 | .02* | -17.33 (-83.35; 48.70) | 33.14 | .83 |
| Cerebral WM volume | -3781.72 (-8228.06; 664.62) | 2231.98 | .30 | 1295.35 (362.62; 2228.08) | 468.21 | .04* | 345.15 (-373.56; 1063.86) | 360.78 | .68 |
| Cerebellar GM volume | 841.30 (-421.41; 2104.00) | 633.85 | .94 | 2.01 (-262.87; 266.90) | 132.97 | 1.00 | -94.22 (-298.32; 109.89) | 102.46 | 1.00 |
| Cerebellar WM volume | 143.25 (-482.01; 768.52) | 313.87 | 1.00 | 3.73 (-127.44; 134.89) | 65.84 | 1.00 | 23.57 (-77.50; 124.64) | 50.73 | 1.00 |
| Ventricular volumes | -634.31 (-1681.77; 413.16) | 525.81 | 1.00 | 67.08 (-152.65; 286.82) | 110.30 | 1.00 | 1.44 (-167.87; 170.75) | 84.99 | 1.00 |
| **Unadjusted for ICV (df = 76)** |  |  |  |  |  |  |  |  |  |
| Cortical volume | 14153.7 (7860.22; 20447.11) | 3159.9 | < .001* | -1882.8 (-3233.29; -532.38) | 678.0 | .02* | 276.6 (-739.25; 1292.35) | 510.0 | .59 |
| Cortical thickness | 0.030 (0.017; 0.044) | 0.007 | < .001* | -0.003 (-0.006; -0.000) | 0.001 | .19 | -0.001 (-0.004; 0.001) | 0.001 | .41 |
| Cortical surface area | 2537.90 (338.26; 4737.48) | 1104.40 | .12 | -309.40 (-781.39; 162.60) | 237.00 | .78 | 209.50 (-145.58; 564.48) | 178.30 | .78 |
| Subcortical volumes | 279.66 (-395.33; 954.64) | 338.91 | .83 | -113.70 (-258.54; 31.14) | 72.72 | .61 | 69.49 (-39.46; 178.44) | 54.70 | .83 |
| Cerebral WM volume | 2325.30 (-5392.42; 10043.12) | 3875.00 | .68 | 1498.50 (-157.57; 3154.60) | 831.50 | .30 | 1362.00 (116.26; 2607.65) | 625.50 | .16 |
| Cerebellar GM volume | 1609.80 (143.58; 3076.02) | 736.17 | .19 | 27.58 (-287.04; 342.20) | 157.97 | 1.00 | 33.74 (-202.92; 270.39) | 118.82 | 1.00 |
| Cerebellar WM volume | 260.68 (-358.42; 879.78) | 310.84 | 1.00 | 7.63 (-125.21; 140.48) | 66.70 | 1.00 | 43.12 (-56.81; 143.05) | 50.17 | 1.00 |
| Ventricular volumes | -222.17 (-1326.55; 882.21) | 554.50 | 1.00 | 80.79 (-156.18; 317.77) | 118.98 | 1.00 | 70.06 (-108.19; 248.31) | 89.50 | 1.00 |

Note: Cortical thickness expressed in millimeters, cortical surface area in millimeters<sup>2</sup>, and remaining variables in millimeters<sup>3</sup>. P-values are adjusted according to a Bonferroni-Holm correction.
\*Statistically significant. <sup>†</sup>Adjusted p-value is < .05, but rounded up to .05. Abbreviations: BMI SDS: Body mass index (kg/m<sup>2</sup>) standard deviation score; CI: Confidence interval; df = Degrees of freedom; GM: Gray matter; ICV: Intracranial volume; SE: Standard error; WM: White matter.

#### Association between BMI and brain morphology

BMI SDS showed a significant positive association with both cortical volume and thickness (Table 2). These were significant in regression models both unadjusted and adjusted for ICV. This indicates that lower BMI (while controlling for age) is associated with reduced relative and absolute cortical volume and thickness. These effects were larger than the effects of age. No other significant associations were evident. As no relationship was found between BMI SDS and cortical surface area, the association between BMI SDS and cortical volume is likely driven by differences in cortical thickness.

#### Interaction effect age * BMI

The interaction term was not significant in any of the models (Table 2), indicating that the influence of age and BMI on brain morphology highlighted above constitute independent effects. Thus, we found no evidence of moderating effects.

Scatterplots depicting the significant associations of BMI SDS and age with brain morphology are presented in Figure 1 (Supplemental Figure S5-6 presents all scatterplots). Note that scatterplots show associations unadjusted for ICV. Inspection of these identified one patient whose cortical volume and thickness estimates were extremely low, and this patient also had the lowest BMI. To rule out the possibility that this patient was driving the observed associations between BMI and brain morphology, we excluded this patient and re-ran the regression models. This did not alter the statistical outcomes of the models. Regression models were also performed after excluding the five male patients, which had no effect on the statistical outcomes of the models.

**Figure 1.**
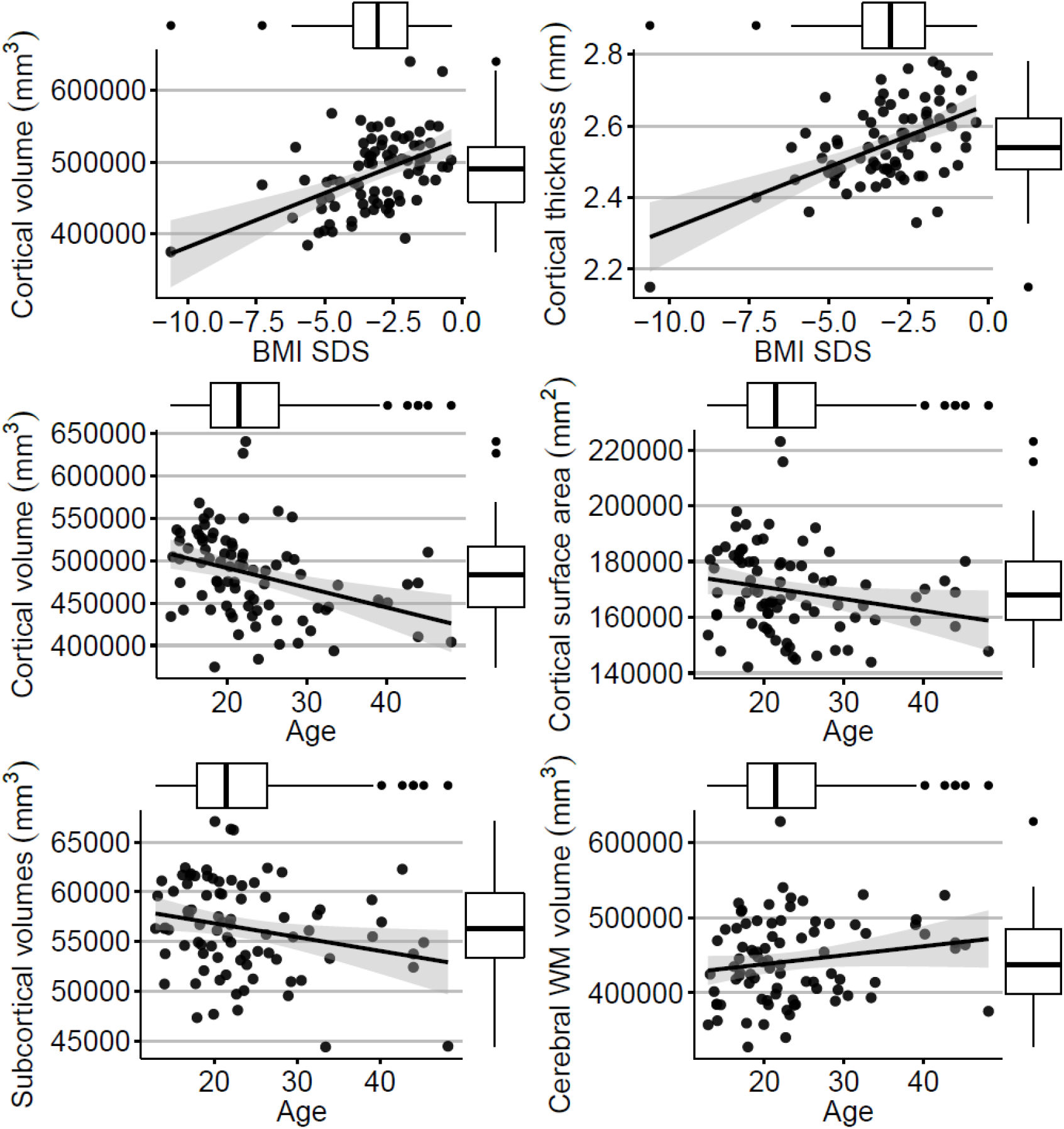
Scatterplots showing associations of BMI SDS and age with brain morphology. Only associations significant in regression models are shown. BMI SDS, Body mass index (kg/m2) standard deviation score; Mm, Millimeters; WM, White matter.

#### 3.3 Correlations between biochemical parameters and brain morphology

Biochemical parameters for most patients were within reference intervals (Supplemental Figure S7-8 present distributions of all biochemical parameters). However, albumin, bicarbonate, pCO_2_, and base-excess were above the reference intervals for a considerable proportion of patients. Additionally, pH was below the reference interval for approximately 50% of patients. Table 3 shows all pairwise partial correlations between biochemical parameters and brain morphology variables. No significant correlations were evident, and the majority of correlation coefficients were of small magnitude. These correlations were also non-significant prior to alpha-level correction. Excluding the five male patients had no effect on the statistical outcomes of the correlations.

**Table 3.** Correlations between biochemical parameters and brain morphology.

|  | Cortical<br>volume | Cortical<br>thickness | Cortical<br>surface area | Subcortical<br>volumes | Cerebral WM<br>volume | Cerebellar GM<br>volume | Cerebellar WM<br>volume | Ventricular<br>volumes |
| --- | --- | --- | --- | --- | --- | --- | --- | --- |
| **Albumin**<br>*n* = 64 | .07<br>*p* = 1.00 | .06<br>*p* = 1.00 | .07<br>*p* = 1.00 | .02<br>*p* = 1.00 | -.02<br>*p* = 1.00 | .11<br>*p* = 1.00 | .09<br>*p* = 1.00 | -.15<br>*p* = 1.00 |
| **CRP**<br>*n* = 64 | .18<br>*p* = 1.00 | .09<br>*p* = 1.00 | .14<br>*p* = 1.00 | .05<br>*p* = 1.00 | -.03<br>*p* = 1.00 | .07<br>*p* = 1.00 | .04<br>*p* = 1.00 | .04<br>*p* = 1.00 |
| **Sodium**<br>*n* = 77 | -.16<br>*p* = 1.00 | -.17<br>*p* = 1.00 | -.08<br>*p* = 1.00 | .17<br>*p* = 1.00 | .05<br>*p* = 1.00 | .21<br>*p* = 1.00 | .13<br>*p* = 1.00 | -.05<br>*p* = 1.00 |
| **Potassium**<br>*n* = 78 | -.04<br>*p* = 1.00 | .08<br>*p* = 1.00 | -.10<br>*p* = 1.00 | .03<br>*p* = 1.00 | -.00<br>*p* = 1.00 | -.17<br>*p* = 1.00 | .02<br>*p* = 1.00 | -.28<br>*p* = .15 |
| **Chloride**<br>*n* = 37 | -.12<br>*p* = 1.00 | .00<br>*p* = 1.00 | -.14<br>*p* = 1.00 | .11<br>*p* = 1.00 | .04<br>*p* = 1.00 | .17<br>*p* = 1.00 | .08<br>*p* = 1.00 | .01<br>*p* = 1.00 |
| **Phosphate**<br>*n* = 78 | .18<br>*p* = 1.00 | .15<br>*p* = 1.00 | .10<br>*p* = 1.00 | -.04<br>*p* = 1.00 | -.06<br>*p* = 1.00 | .14<br>*p* = 1.00 | .15<br>*p* = 1.00 | .01<br>*p* = 1.00 |
| **Calcium**<br>*n* = 67 | .03<br>*p* = 1.00 | -.17<br>*p* = 1.00 | .20<br>*p* = 1.00 | -.04<br>*p* = 1.00 | .00<br>*p* = 1.00 | .23<br>*p* = 1.00 | .05<br>*p* = 1.00 | -.05<br>*p* = 1.00 |
| **Calcium corrected<sup>†</sup>**<br>*n* = 53 | -.16<br>*p* = 1.00 | -.36<br>*p* = .14 | .08<br>*p* = 1.00 | -.03<br>*p* = 1.00 | -.00<br>*p* = 1.00 | -.07<br>*p* = 1.00 | -.13<br>*p* = 1.00 | .16<br>*p* = 1.00 |
| **Hemoglobin**<br>*n* = 65 | .18<br>*p* = 1.00 | .23<br>*p* = .98 | -.01<br>*p* = 1.00 | -.08<br>*p* = 1.00 | -.26<br>*p* = .60 | .18<br>*p* = 1.00 | -.04<br>*p* = 1.00 | .24<br>*p* = .65 |
| **EVF**<br>*n* = 54 | .14<br>*p* = 1.00 | .21<br>*p* = 1.00 | -.06<br>*p* = 1.00 | -.07<br>*p* = 1.00 | -.24<br>*p* = 1.00 | .23<br>*p* = 1.00 | -.08<br>*p* = 1.00 | .29<br>*p* = .56 |
| **LD**<br>*n* = 55 | -.10<br>*p* = 1.00 | -.16<br>*p* = 1.00 | .05<br>*p* = 1.00 | -.29<br>*p* = .45 | .00<br>*p* = 1.00 | -.03<br>*p* = 1.00 | -.14<br>*p* = 1.00 | .02<br>*p* = 1.00 |
| **pH**<br>*n* = 28 | .11<br>*p* = 1.00 | .13<br>*p* = 1.00 | .03<br>*p* = 1.00 | -.12<br>*p* = 1.00 | -.30<br>*p* = 1.00 | .11<br>*p* = 1.00 | -.09<br>*p* = 1.00 | .13<br>*p* = 1.00 |
| **pCO<sub>2</sub>**<br>*n* = 28 | -.18<br>*p* = 1.00 | -.17<br>*p* = 1.00 | .11<br>*p* = 1.00 | -.16<br>*p* = 1.00 | .22<br>*p* = 1.00 | .07<br>*p* = 1.00 | .18<br>*p* = 1.00 | -.38<br>*p* = .65 |
| **Bicarbonate**<br>*n* = 29 | -.13<br>*p* = 1.00 | -.12<br>*p* = 1.00 | -.10<br>*p* = 1.00 | -.25<br>*p* = 1.00 | .08<br>*p* = 1.00 | .16<br>*p* = 1.00 | .17<br>*p* = 1.00 | -.34<br>*p* = .77 |
| **Base-excess**<br>*n* = 30 | -.17<br>*p* = 1.00 | -.11<br>*p* = 1.00 | -.15<br>*p* = 1.00 | -.30<br>*p* = 1.00 | .01<br>*p* = 1.00 | .23<br>*p* = 1.00 | .16<br>*p* = 1.00 | -.30<br>*p* = 1.00 |

Note: P-values are adjusted according to a Bonferroni-Holm correction, and all correlations are non-significant. <sup>†</sup>Calcium corrected for albumin levels. *Abbreviations:* GM, Gray matter; WM, White matter.

## 4. DISCUSSION

The current study showed that age and BMI are independently associated with brain morphology in AN. Age-related effects were observed for several measures of brain morphology, and were generally concordant with prior studies of typical neurodevelopment. Lower BMI was associated with greater reductions to cortical volume and thickness, indicating a relationship between severity of emaciation and brain alterations. However, we did not find evidence to support an interaction effect between age and BMI. Moreover, none of the biochemical parameters were associated with brain morphology.

Our finding that increasing age is associated with decreased relative cortical and subcortical volumes, decreased cortical surface area, and increased cerebral WM volume align with prior studies of typical neurodevelopment (Lebel & Beaulieu, 2011; Mills et al., 2016; Tamnes et al., 2017). In the models on absolute brain morphology, only cortical volume was associated with age, which may indicate that including ICV as a nuisance variable may better capture the age-effects in heterogeneous AN samples. These results generally indicate typical neurodevelopmental patterns among patients with AN. A possible exception was observed for cortical thickness, as we did not find a significant age related association, while cortical thickness is known to decrease with increasing age from childhood to adulthood (Tamnes et al., 2017). However, a negative association was found prior to alpha-level correction, which may indicate insufficient statistical power. Interestingly, however, a previous study of patients with AN also failed to find a relationship between age and cortical thickness (King et al., 2015), which the authors suggested may be related to interrupted neurodevelopment. Another study (Kaufmann et al., 2020) reported that age correlated with restoration of cortical thickness during treatment, with younger patients recuperating faster than older patients. This may reflect increased brain plasticity among younger patients, and together with our findings highlights the need for future longitudinal studies to consider individual development of brain morphology in AN. One caveat of our study is that we were unable to disentangle age effects from duration of AN, but prior studies have reported that duration of AN is unrelated to brain morphology (Bernardoni et al., 2016; Kaufmann et al., 2020; Nickel et al., 2018).

Lower BMI was associated with decreased relative and absolute cortical volume and thickness, indicating that severity of emaciation is related to brain mass reductions. These associations corresponded to 0.03 millimeter decrease in cortical thickness for each unit decrease in BMI SDS, and 0.1 deciliter decrease in cortical volume for each unit decrease in BMI SDS (relative to ICV). The effects of BMI were larger than the effects of age, in line with prior reports (Bernardoni et al., 2016). Previous studies have produced inconsistent results regarding whether severity of emaciation is significantly associated with brain morphology; with some supporting this association (Lavagnino et al., 2016; Nickel et al., 2018) while others do not (Curzio et al., 2020; Fuglset et al., 2016; King et al., 2015). The reasons for this discrepancy are unclear, though statistical power may constitute one important factor. Our study included a large sample of patients who varied considerably in BMI, which may have provided sufficient statistical power. Our findings align with converging evidence from longitudinal studies of the strong influence of weight restoration on cortical volume (Bernardoni et al., 2016; Kaufmann et al., 2020; Mainz et al., 2012; Roberto et al., 2011) and thickness (Bernardoni et al., 2016; Kaufmann et al., 2020). Of note, we focused on global measures of brain morphology and were unable to investigate associations between BMI and local measures of brain morphology. Some studies have reported increased local brain volume and cortical thickness among individuals with AN (Frank et al., 2013; Lavagnino et al., 2018; Leppanen, Sedgewick, Cardi, Treasure, & Tchanturia, 2019), raising the possibility of negative associations between BMI and local measures of brain morphology that we were unable to characterize.

In contrast, we found no evidence of an association between BMI and other measures of brain morphology, including subcortical volumes and WM volumes. Reductions of these volumes are well documented in AN (Bernardoni et al., 2016; Miles et al., 2018; Titova et al., 2013). One possible reason for these null-findings is that severity of emaciation is not as directly related to these tissues. We also showed that BMI was significantly associated with cortical thickness but not cortical surface area. Few studies have investigated cortical surface area in AN and current findings are inconsistent; with one study reporting similar cortical surface area (Miles et al., 2018) and another reporting decreased surface area (Leppanen et al., 2019) as compared to healthy individuals. As cortical volume is defined by both cortical thickness and surface area, our findings suggest that the association between BMI and cortical volume is primarily due to thinning of the cortex, and not decreased cortical surface area.

While our findings highlight independent associations of both age and BMI with brain morphology, no significant interaction effect was evident. Thus, there was no evidence of a moderating role of age on the association between BMI and brain morphology. Some have noted that younger individuals with AN may have particularly smaller brain volumes (Seitz et al., 2016), raising the possibility that malnutrition and low body weight influences adolescents more than adults. The current study showed no such relation. As only a quarter of patients in our sample were below 18 years of age, it is possible we had insufficient statistical power to reveal such effects.

We attempted to elucidate the mechanisms underlying the covariation between BMI and brain morphology by examining potential associations between various biochemical parameters and brain morphology. Despite that several of the parameters were outside reference intervals for many patients (indicative of acid-base imbalance), none of the correlations with brain morphometry measures were significant, and the magnitudes of the correlations were generally small. This indicates that the mechanisms underlying reductions in brain mass may be unrelated to the biochemical parameters we considered. Few studies have examined similar relationships between biochemical parameters and brain morphology in patients with AN. One study (Bernardoni et al., 2016) considered serum albumin and specific gravity of urine, but found that these were within normal range and thus unlikely to contribute to observed changes to brain morphology. We similarly failed to detect an association between albumin and brain morphology, despite albumin levels being high for many patients. This may suggest that dehydration and fluid shifts are unlikely causes of the brain mass reductions. However, our null-findings may be due to insufficient statistical power, as information on biochemical parameters was unavailable for many patients. It is also possible that brain morphology alterations in AN are influenced by other biochemical and hormonal parameters not considered in our study. For example, previous studies have found an association between brain morphology and cortisol (Castro-Fornieles et al., 2009; Mainz et al., 2012) and follicle-stimulating hormone (Mainz et al., 2012). Future highly powered studies are needed for continued investigation of the underlying mechanisms of brain mass reductions and normalization in AN.

Our study had several limitations, most of which are related to the clinical nature of our data. Collection of neuroimaging data was not performed in a systematic manner, and was attained on several different scanners. Due to this heterogeneity, we were unable to control for scanner and sequence related variability. For these reasons, we focused on global brain morphology, which precluded us from investigating local morphology measures. Patients were also in different stages of treatment and weight restoration, but rate of weight gain was slow and most were still considerably underweight at the time of the MRI examination. This natural variability in BMI also constitutes a strength of the study, as we were able to sample patients with varying degrees of emaciation. Moreover, as our study was without a comparison group, we cannot determine the extent to which our patients were characterized by brain mass reductions relative to healthy individuals. However, such reductions are well-documented in previous studies, and our findings highlight the influence of BMI on brain morphometry. Finally, the cross-sectional study design is a sub-optimal and indirect way of investigating maturation and development, and we therefore cannot exclude the possibility that age and brain morphology related findings reflect factors other than neurodevelopment (e.g. AN duration).

In conclusion, our study highlights that both age and BMI are independently associated with variations in brain morphology in AN. However, there was no evidence of an interaction effect between age and BMI, and thus no indication that age moderates the relationship between BMI and brain morphology. These results suggest the presence of at least partial normal neurodevelopmental patterns among individuals with AN. Importantly, our findings also showed that severity of emaciation is related to brain morphology reductions, underscoring the importance of weight restoration. We did not detect associations between biochemical parameters and brain morphology, and further studies are needed to shed light on potential mechanisms underlying the brain alterations. Our findings align with prior work highlighting the positive message that weight gain leads to normalization of brain morphology. Communicating this to patients and their families may provide hope and reassurance.

## Data Availability

Due to issues regarding confidentiality and ethics, data cannot be shared.

## Acknowledgments

LB is funded by the South-Eastern Norway Regional Health Authority (#2017083). CKT is funded by the Research Council of Norway (#288083, #223273). CKT and LBN are funded by the South-Eastern Norway Regional Health Authority (#2019069).

